# Reliability of force plate-based measures of standing balance in the sub-acute stage of post-stroke recovery

**DOI:** 10.1101/2023.05.18.23290052

**Authors:** Raabeae Aryan, Elizabeth Inness, Kara K. Patterson, George Mochizuki, Avril Mansfield

## Abstract

**Background:** Difficulty controlling balance is one of the major contributors to the increased risk of falls among individuals with stroke. It is important to use reliable and objective measures to evaluate balance impairments post-stroke to inform more effective post-stroke rehabilitation.

**Objectives:** To examine the relative and absolute reliabilities of force plate-based balance measures in quiet standing, in the sub-acute stage of stroke recovery.

**Methods:** Twenty-four people with sub-acute stroke (mean age=61 years) performed two trials of quiet standing, each 30 seconds long. Sixteen force plate-based balance measures in the time, frequency, or nonlinear domains were calculated. Within-session test-retest reliabilities were investigated using intraclass correlation coefficient (ICC), standard error of measurement, and minimal detectable change.

**Results:** Mean speed of displacements of the centre of pressure along the anterior-posterior axis (ICC=0.91; CI_95%_=[0.83, 0.95]), and directional weight-bearing asymmetry (ICC=0.91; CI_95%_=[0.82, 0.95]) demonstrated high relative reliabilities, followed by the speed-based symmetry index and absolute weight-bearing asymmetry (both ICCs=0.86; CI_95%_=[0.74, 0.93]).

**Conclusions:** Mean speeds of centre of pressure, directional weight-bearing asymmetry, and speed-based symmetry index are the most reliable measures that can be included in the balance assessments of individuals within the sub-acute stage of post-stroke recovery. These findings can better inform clinicians about the specific balance problems experienced by people in this population.

## 1. INTRODUCTION

Poor balance control, while standing or moving, is one of the main contributors to the increased risk of falls among people with stroke [1–3]. Falls can potentially limit an individual’s capacity to participate in daily life activities, lead to physical injuries, and can contribute to decreased quality of life [3,4]. Thus, precise and objective examination of an individual’s balance control is a crucial step for implementing effective post-stroke rehabilitation.

Performance-based balance measures are frequently used to reflect a patient’s balance ability [5]. However, it is likely that therapists use their observation of a patient’s performance on individual items of these scales, rather than the score itself, to guide clinical decision-making [6,7]. In contrast, instrumented balance assessments, such as force plate-based approaches, are used to provide more objective insights into the specific balance deficits experienced by people with stroke [8]. Force plates can be used to measure fluctuations of the centre of pressure (COP) under both feet combined when standing still [9]. Two adjacent force plates can also be used to determine the extent of contributions of individual limbs to balance control, synchrony between the lower limbs, and asymmetric weight distribution. These measures are especially important in post-stroke balance assessments, where the asymmetric motor impairments exist [6]. However, despite the promising ability of force plate-based measures in detecting underlying mechanisms of impaired balance control, little is known about their reliability for assessing balance among people with sub-acute stroke.

Although previous studies have reported reliability of force plate-based measures of standing balance in different clinical populations [10,11], only few have reported reliability of these measures in stroke, either in the chronic stage [12,13], or collectively across multiple stages of post-stroke recovery [14,15]. Few studies have established reliability of force plate-based measures of standing balance in the sub-acute stage of stroke recovery. The sub-acute stage of stroke recovery (7 days-6 months post-stroke [16]) is a critical period for neuroplasticity and spontaneous recovery [16], in which patients experience rapid changes in their physiological and functional status. Only Gray *et al*. (2014) has directly focused on the reliability of some time-domain force plate-based measures of standing balance in sub-acute stroke [17], and found that area of COP displacements and mean speed of COP were the two most reliable measures during this post-stroke stage of recovery [17].

Time-domain force plate-based measures can provide clinicians with useful information about asymmetry of balance control, status of between-limbs coordination, and forces required to regulate postural sway [6,18–21]. Alternatively, frequency and nonlinear domains of force plate-based measures can be useful for investigating contributions of different sensory systems to balance control [22,23], and the regularity of COP time-series [24], respectively. While force plate-based frequency-domain [22] and nonlinear-domain [24] balance measures have been used to detect balance deficits, no previous studies have attempted to establish their reliability in the sub-acute stage of post-stroke recovery.

Given the paucity of research investigating reliability of force plate-based measures that are believed to be useful in evaluation of post-stroke balance control, the primary objective of our study was to determine within-session relative and absolute reliabilities of force plate-based time, frequency, and nonlinear-domain balance measures of quiet standing in people within the sub-acute stage of stroke recovery. Our main goal was to identify the balance measures with high reliability, among the measures that are believed to be appropriate for use in post-stroke balance assessments and rehabilitation. We hypothesized that our select set of force plate-based measures would demonstrate clinically acceptable levels of relative reliability (ICC>0.8) [25,26] and low measurement error.

## 2. METHODS

### 2.1. Participants

This study was a retrospective secondary analysis of the force plate data collected for two other larger projects [27,28]. Both original studies received approvals from the institution’s Research Ethics Board. Participants in the original studies were patients (either post-stroke or acquired brain injury) and healthy adults, who were recruited between August 2013 and January 2017. Written informed consent was obtained from the eligible individuals. Common inclusion criteria of the original studies were: 1) age≥18 years old; 2) ability to stand independently for one minute; and 3) ability to understand test instructions in English.

Participants were included in the present analysis if they: 1) were individuals with stroke who were receiving or had recently completed inpatient post-stroke rehabilitation at the Toronto Rehabilitation Institute (four participants completed assessments 26-38 days post-discharge from inpatient rehabilitation, while they were still in the sub-acute stage); 2) were in the sub-acute stage of stroke recovery at the time of the force plate assessments; and 3) had two force plate-based assessments of quiet standing balance in one day, each lasting at least 30 seconds. We excluded participants if they had any condition besides stroke that could potentially affect their balance control (e.g., other neurological conditions, amputations). From a total of 115 participants in the original studies, we included 24 participants who met all criteria of the present study.

### 2.2. Assessment procedures

Participants were instructed to stand in a standard foot position on two adjacent force plates (OR6-7-2000, Advanced Medical Technology Inc., Watertown, Massachusetts, USA), with each foot placed on a separate force plate and oriented 14 degrees outwards, while centres of their heels were 17 cm apart [29]. Participants stood quietly for 30 seconds in their preferred weight-distributed stance position, with their eyes open, and arms by their sides (if able), while wearing comfortable shoes. Participants who typically wore an ankle-foot orthosis kept their orthosis on during data acquisition. To ensure safety during the force plate-based assessments, participants were supervised by a research assistant and a physiotherapist.

Two quiet standing trials were performed in one session (one test trial, one retest trial). The time-intervals between test and retest varied among the participants; however, test and retest trials were not closer than 10 seconds apart. Participants could request rest breaks if needed during the assessment sessions. Force plate data were sampled at 256 Hz and stored for offline processing. Individuals’ height, weight, age, lesion side(s), paretic side, and Berg Balance Scale scores [5] were collected (either directly from participants or from their hospital charts) or assessed by the same research assistant/or physiotherapist. In two cases, where the participants had bilateral stroke lesions, the more-affected side of their body was determined from information in participants’ medical charts and/or performance-based assessments.

### 2.3. Data processing

Forces and moments were filtered using a low-pass 4^th^-order zero phase-lag Butterworth filter at 10 Hz. COP under each foot separately, and under both feet combined (net-COP) were calculated for each trial, along the anterior-posterior (AP) and medial-lateral (ML) directions.

COP time-series’ were down-sampled to 64 Hz; and their means were subtracted from the signals prior to calculating balance measures in the time, frequency, and nonlinear domains. Down-sampling by a factor of 4 allowed us to prepare the COP time-series for computing our nonlinear balance measure (i.e., sample entropy) and its *a priori* parameters (i.e., segment length, and tolerance radius) [30], while staying above the suggested optimal minimum sampling frequency to calculate the remainder of our force-based balance measures [31]. All measures were computed offline, using a custom-written MATLAB routine (MATLAB and Statistics Toolbox Release 2016b, The MathWorks, Inc., Natick, Massachusetts, USA). To compute the frequency-domain measures, the first 5 seconds from the COP time-series’ were removed, and MATLAB Welch’s power spectral estimation function (segment length 800 points, with 25% overlap, frequency resolution of 0.08 Hz) was used. Thus, frequency-domain analysis was performed on the remaining 25 seconds, on the frequency band of 0-10 Hz. All other measures were calculated from the full 30-second trial.

#### 2.3.1. Time-domain measures

The following measures were selected according to the recommendations regarding the use of force plate-based measures [6,9,32]:

a. Root mean square (RMS) of displacements of the net-COP along the AP and ML axes: RMS of COP reflects the variability of the time-series [6,33], and is often used as a proxy to quantify amplitude of postural sway [34], since RMS will generally increase as sway increases [6].
b. Mean speeds of net-COP along the AP, and ML directions: Speed of COP represents the neuromuscular control exerted by the muscles in the lower extremities to respond to postural sway [20,33]; higher speeds can be indicative of a poorer balance control [23,33].
c. The 95% ellipse area of the net-COP (both feet combined) was calculated based on the principal component analysis approach [35]. Reportedly, the smaller the ellipse area, the better the general performance of the balance control system [32].
d. Directional, and absolute weight-bearing asymmetries (dir-WBA and abs-WBA): WBA addresses the imbalance in weight distribution between the lower extremities of individuals with hemiparesis while standing in their preferred quite stance posture [6]. Dir-WBA and abs-WBA were calculated as follows [6]:

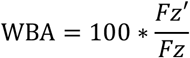

To yield the dir-WBA, *FZ*′ is the mean vertical ground reaction force beneath the paretic limb. To calculate the abs-WBA, *FZ*′ is the absolute difference between the mean vertical ground reaction forces under both feet. *FZ* represents the total mean vertical ground reaction force beneath both feet.
e. Symmetry indices were calculated for both the displacement and speed of COP. We calculated displacement-based and speed-based symmetry indices using the following formula, in which the *RMS* is the RMS of displacement or speed of the AP-COP under each feet [6,36]:

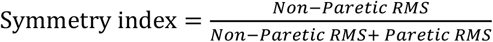 Symmetry index is reportedly a clinically useful measure that quantifies individual-limb contributions to balance control [6].
f. Inter-limb cross-correlation reflects how both lower extremities work in synchrony while standing [6], and was calculated by measuring the correlation of displacements of the AP-COP time series between the paretic and non-paretic extremities in quiet standing [37].

#### 2.3.2. Frequency-domain measures

We selected mean and median power frequencies of the AP and ML components of the net-COP time-series, since these measures have been previously used to characterize quiet standing balance in stroke and other populations [22,38,39]. Studying the power spectrum of COP time-series can be useful to demonstrate the relative contributions of different sensory systems to balance control. Higher frequencies indicate faster and smaller balance adjustments [9,33], which are possibly associated with fast corrections in response to sudden instabilities [19] and can be indicators of increased contribution of the somatosensory system to balance control [40].

#### 2.3.3. Nonlinear-domain measures

Sample entropy of COP is a nonlinear-domain measure which is thought to reflect the attentional demand for balance control [24]; thus, it can highlight the regularity of COP time-series and complexity of underlying balance control processes. We calculated sample entropy using MATLAB code available on PhysioNet [41]. The net-COP time-series’ were detrended and normalized by the standard deviation of the signal within the code. According to methods described elsewhere [42], we determined the optimal values for *m* (segment length) and *r* (tolerance radius) as *m*=2, *r*=0.03, and *m*=2, *r*=0.02 for net AP and ML-COP displacements, respectively.

### 2.4. Statistical analysis

To detect any bias between repeated measurements, paired t-tests were used to compare force plate-based measures between the test and retest assessments. Bland-Altman plots were generated to visualize biases and agreements between the test and retest trials. Within-session test-retest relative reliability of the force plate-based measures of standing balance was calculated based on a two-way mixed effect ANOVA model to demonstrate agreement between repeated measurements (ICC _2,1_) [43]. ICC values<0.5, between 0.5 and 0.75, between 0.75 and 0.9, and >0.9 were considered as poor, moderate, good, and excellent relative reliability, respectively [43].

Absolute reliability was determined using the standard error of measurement (SEM), and minimal detectable change (MDC). SEM demonstrates a change in the measurement score which is due to a random error [44]. SEM was calculated using the following formula, where *SD* is the standard deviation of all test and retest assessments [45]: 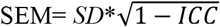.

MDC is the smallest change in the test score that is not due to a random error and represents a true change [44]. MDC was calculated as follows: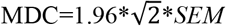. Statistical significance level was considered as α=0.05. Statistical analysis was completed using the R 3.3.2 (R Core Team, 2017).

## 3. RESULTS

Demographic characteristics of participants are presented in Table 1. The average test-retest time interval was 11.5 minutes (standard deviation=8.9 minutes; range=10 seconds to 27.5 minutes; test-retest intervals for five participants were≤1 minute). Mean, standard deviation, and p-values of comparison analysis of repeated measurements are presented in Table 2. There were no statistically significant differences between the values of force plate-based measures in test and retest assessments (p-value>0.05). Relative (ICC_2,1_ and 95% confidence intervals, CI) and absolute reliability (SEMs, 95% MDCs) values are presented in Table 2. Considering the 95% confidence intervals for each ICC value, mean speed of AP-COP, and dir-WBA demonstrated good-to-excellent relative reliability (both ICCs=0.91; 95% CI=0.82-0.95). Speed-based symmetry index (ICC=0.86; 95% CI=0.74-0.93), abs-WBA (ICC=0.86; 95% CI=0.74-0.93), and mean speed of ML-COP (ICC=0.82; 95% CI=0.68-0.91) demonstrated moderate-to-excellent relative reliability. Among the frequency-domain measures, mean power frequency of AP-COP demonstrated moderate-to-good relative reliability (ICC=0.79; 95% CI=0.62-0.89). For nonlinear-domain measures, sample entropy of AP-COP also demonstrated moderate-to-good relative reliability (ICC=0.79; 95% CI=0.63-0.89). The lowest relative reliability values, among all categories, were observed for the RMS of AP-COP (ICC=0.41) and median power frequency of AP-COP (ICC=0.44). Dir-WBA demonstrated high absolute reliability (SEM=2.53% body weight, MDC=7.02% body weight); while the lowest absolute reliability value was observed for the ellipse area (SEM=155.7 mm^2^, MDC=431.6 mm^2^). Bland-Altman plots demonstrated no biases for mean speeds of AP-COP and ML-COP, dir-WBA, and speed-based symmetry index (Figures 1 and 2).

**Table 1:** Participant characteristics. Values presented are means with standard deviations in parentheses for continuous variables, and counts for categorical variables.

| **Participant characteristics** |  |
| --- | --- |
| **Sex (number)** |  |
| Female | 6 |
| Male | 18 |
| **Age (years)** | 61.1 (12.8) |
| Range | 33.0-83.0 |
| **Height (cm)** | 171.1 (8.8) |
| Range | 158.0-191.0 |
| **Mass (kg)** | 79.0 (16.7) |
| Range | 54.1-116.8 |
| **Stroke type** |  |
| Ischemic | 16 |
| Haemorrhagic | 7 |
| Unknown | 1 |
| **Affected side of the brain (number)** |  |
| Right | 10 |
| Left | 12 |
| Both | 2 |
| **More-affected side of the body (number)** |  |
| Right | 12 |
| Left | 12 |
| **Time post-stroke (days)** | 41.0 (21.9) |
| Range | 12-95 |
| **Berg Balance Scale (score)** | 46.3 (11.6) |
| Range | 6-56 |

**Table 2:** Means (and standard deviations), p-values of paired comparisons of test and retest assessments, and reliability values of force plate-based measures of standing balance, derived from the net-COP time-series.

| Measures | Test | Retest | p | ICC<sub>2,1</sub> | CI<sub>95%</sub> | SEM | MDC |
| --- | --- | --- | --- | --- | --- | --- | --- |
| RMS AP-COP (mm) | 6.1 (2.3) | 6.7 (2.7) | 0.21 | 0.41 | [0.10, 0.65] | 1.89 | 5.23 |
| RMS ML-COP (mm) | 4.0 (1.9) | 4.6 (2.2) | 0.07 | 0.74 | [0.54, 0.86] | 1.05 | 2.91 |
| 95% Ellipse area (mm<sup>2</sup>) | 287 (200) | 359 (256) | 0.12 | 0.54 | [0.26, 0.74] | 155.7 | 431.6 |
| dir-WBA (body weight%) | 45.9 (8.2) | 45.3 (8.7) | 0.42 | 0.91 | [0.82, 0.95] | 2.53 | 7.02 |
| abs-WBA (body weight%) | 13.2 (12.6) | 14.5 (13.2) | 0.36 | 0.86 | [0.74, 0.93] | 4.78 | 13.3 |
| Inter-limb cross-correlation | 0.75 (0.29) | 0.76 (0.31) | 0.88 | 0.70 | [0.48, 0.84] | 0.16 | 0.45 |
| Symmetry index, displacement | 0.54 (0.12) | 0.57 (0.11) | 0.12 | 0.70 | [0.49, 0.84] | 0.06 | 0.17 |
| Symmetry index, speed | 0.59 (0.12) | 0.60 (0.11) | 0.38 | 0.86 | [0.74, 0.93] | 0.04 | 0.12 |
| Mean speed AP-COP (mm/s) | 14.0 (8.2) | 15.1 (10.5) | 0.17 | 0.91 | [0.83, 0.95] | 2.83 | 7.84 |
| Mean speed ML-COP (mm/s) | 8.2 (3.2) | 8.7 (4.2) | 0.24 | 0.82 | [0.68, 0.91] | 1.59 | 4.41 |
| Mean power freq. AP-COP (Hz) | 0.33 (0.19) | 0.33 (0.21) | 0.95 | 0.79 | [0.62, 0.89] | 0.09 | 0.25 |
| Mean power freq. ML-COP (Hz) | 0.36 (0.14) | 0.34 (0.13) | 0.26 | 0.75 | [0.55, 0.86] | 0.07 | 0.19 |
| Median power freq. AP-COP (Hz) | 0.23 (0.07) | 0.22 (0.10) | 0.86 | 0.44 | [0.12, 0.67] | 0.06 | 0.18 |
| Median power freq. ML-COP (Hz) | 0.30 (0.15) | 0.28 (0.16) | 0.40 | 0.53 | [0.24, 0.73] | 0.09 | 0.26 |
| Sample entropy AP-COP (bits) | 0.96 (0.43) | 0.90 (0.48) | 0.37 | 0.79 | [0.63, 0.89] | 0.21 | 0.57 |
| Sample entropy ML-COP (bits) | 1.19 (0.37) | 1.09 (0.36) | 0.06 | 0.72 | [0.50, 0.85] | 0.19 | 0.54 |

COP: net centre of pressure. AP: anterior-posterior. ML: medial-lateral. dir-WBA: directional weight-bearing asymmetry. abs-WBA: absolute weight-bearing asymmetry. RMS: root mean square. Freq: frequency. ICC: intraclass correlation coefficient. CI: confidence intervals of ICC. p: p-value. SEM: standard error of measurement. MDC: 95% minimal detectable change.

**Figure 1:**
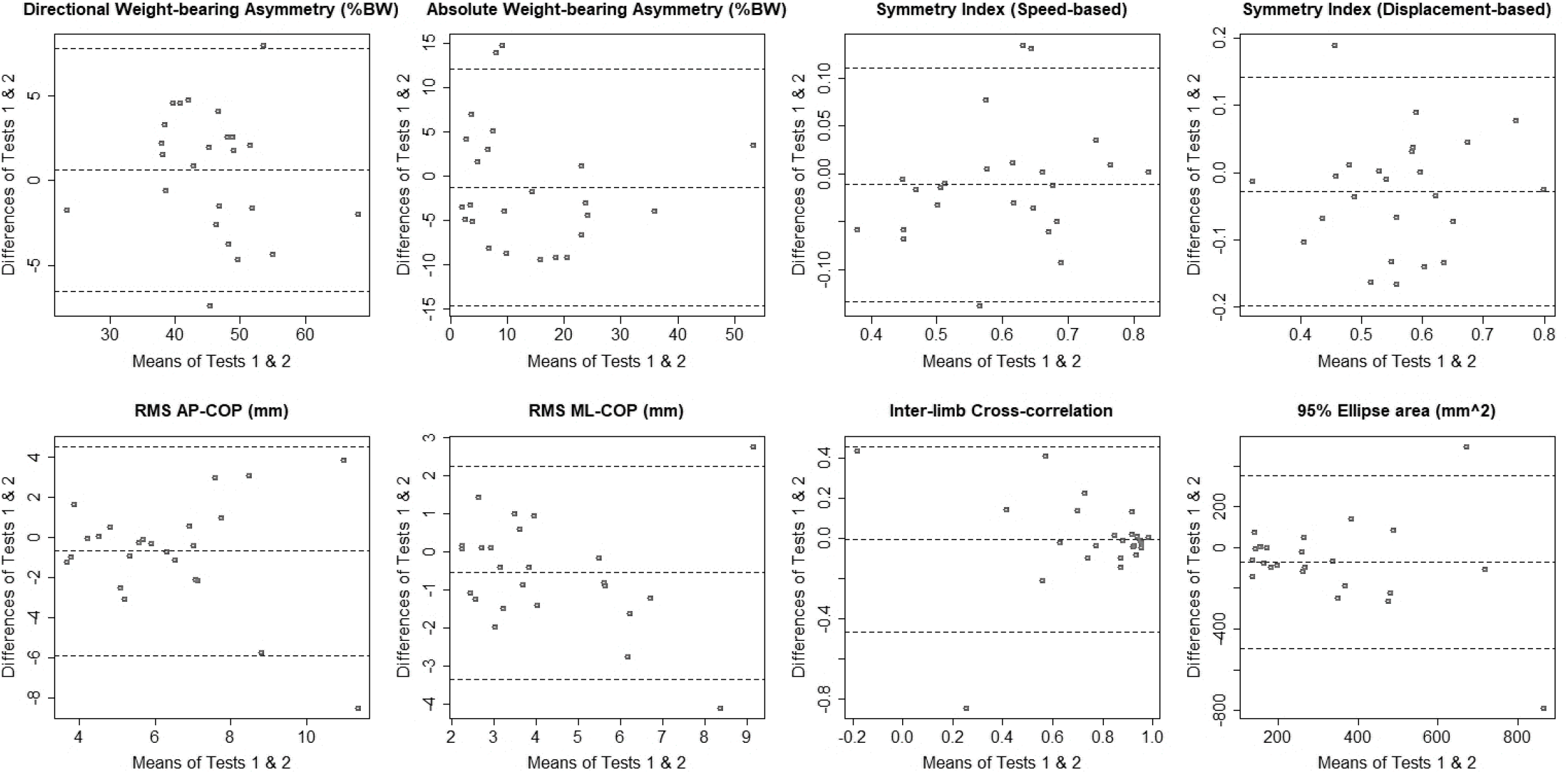
Bland-Altman plots of agreements between test and retest assessments. Each plot depicts the mean of both test and retest (x-axis) against the difference of test and retest assessment (y-axis).

**Figure 2:**
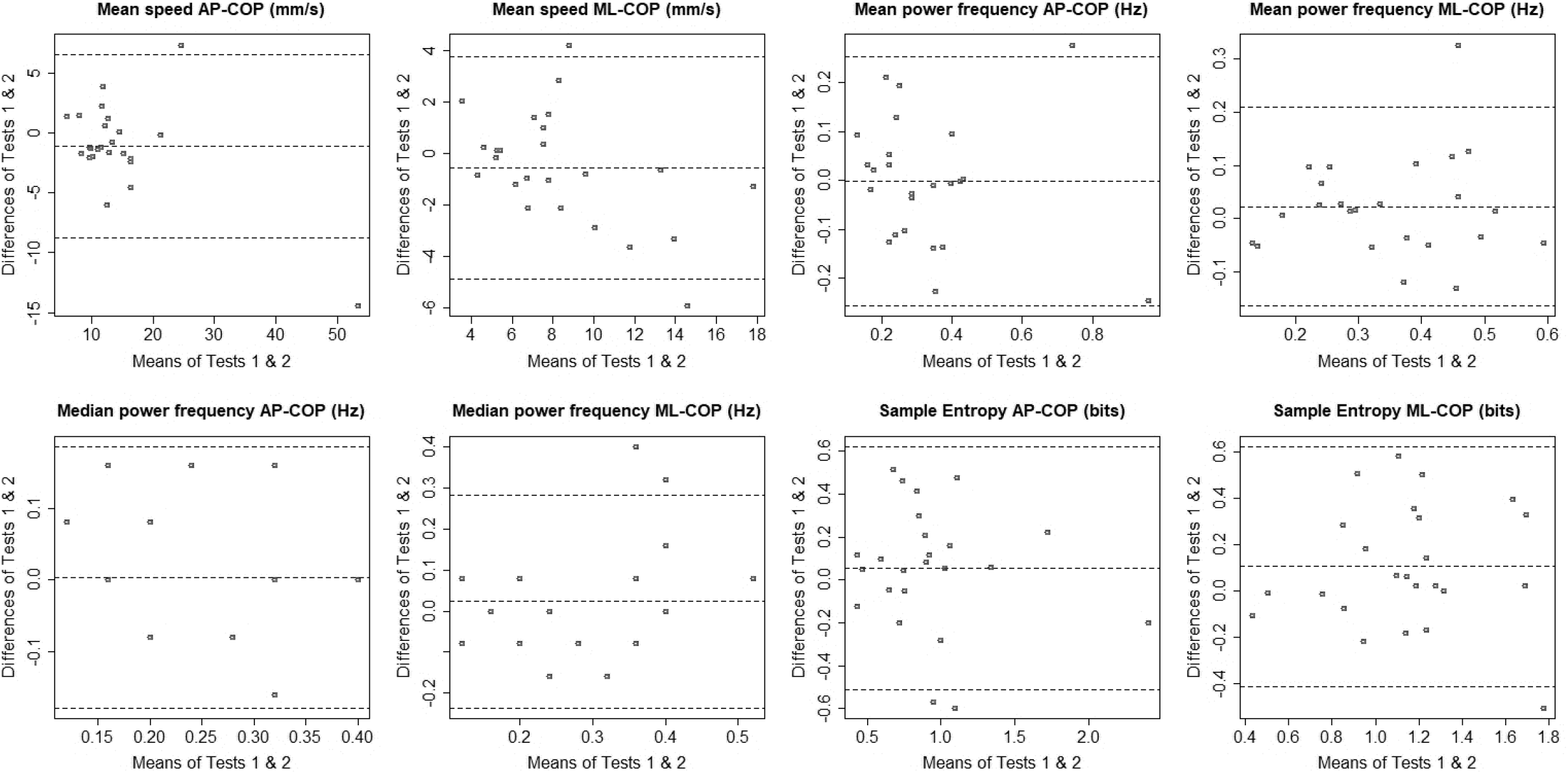
Bland-Altman plots of agreements between test and retest assessments. Each plot depicts the mean of both test and retest (x-axis) against the difference of test and retest assessment (y-axis).

## 4. DISCUSSION

The primary objective of this study was to establish the relative and absolute reliabilities of the force plate-based measures of balance, derived from one trial of 30 seconds-long quiet standing, in individuals assessed during the sub-acute stage of stroke recovery. Findings of this study demonstrated a range of reliability values across all three categories of force plate-based balance measures. Overall, the mean speed of AP-COP and dir-WBA showed high relative reliability, followed by the mean speed of ML-COP and speed-based symmetry index. Dir-WBA also had high absolute reliability for use in the sub-acute stage of stroke recovery.

Speed of COP has been previously reported as a highly reliable force plate-based balance measure in older adults [47]. Reliability values of the speeds of AP-COP and ML-COP in our work are approximately in line with the reliability of the total speed of COP under the paretic and non-paretic lower limbs in a previous study in sub-acute stroke [17]. This previous study found that speeds of COP under both paretic and non-paretic limbs had good relative and absolute reliabilities (ICC=0.82).

Dir-WBA had both high relative reliability and absolute reliability (low measurement error); this finding is in accordance with previous work performed across multiple stages of stroke recovery [15], which reported an excellent ICC estimate value for WBA (ICC≥0.95). Their slightly higher reliability value for WBA can be partly due to using average of multiple trials, and/or including individuals with chronic stroke who may have less variable weight distribution. Abs-WBA, on the other hand, had an ICC estimate value of 0.86, while having high measurement error (high SEM). This high measurement error may have occurred because abs-WBA shows the absolute difference of the proportion of weight distribution between the paretic and non-paretic sides; that is, one-percent change on each lower extremity will be reflected as a two-percent change of the abs-WBA. This can be a possible reason that has amplified the measurement error of the abs-WBA and made it highly variable during test and retest assessments, compared to the dir-WBA. Therefore, use of the dir-WBA is suggested over the abs-WBA, as it has a higher relative reliability (ICC estimate>0.9) and low measurement error. Dir-WBA is also more informative clinically, because it addresses both the direction and magnitude of asymmetric weight-bearing.

We showed that 95% ellipse area, given the confidence intervals of its ICC, has poor-to-moderate relative reliability (95% CI=0.26-0.74), whereas Gasq *et al*. demonstrated good reliability for this measure (ICC=0.76) [14]. This difference can be in part due to Gasq *et al*. using a longer COP signal than us (approximately 51 seconds), and taking the average of multiple trials. It has been shown that reliability of force plate measures is improved when the sampling duration is increased and/or when multiple trials are averaged [48]. Moreover, in our study, ellipse area had high measurement error (48% of its mean). Again, this emphasizes the high within-subjects variations of the ellipse area calculated from the net-COP time-series; this is likely due to the lack of sufficient control of the COP under the paretic side and its effects on the net-COP values.

We found poor-to-good relative reliability (given the ICC confidence intervals) for the displacement-based symmetry index, whereas the speed-based symmetry index showed moderate-to-excellent relative reliability. These results are in accordance with another study in chronic stroke, in which a slightly higher reliability was shown for speed-based symmetry index than the displacement-based index [13]. It is speculated that the displacement-based symmetry index shows lower relative reliability than the speed-based index because the RMS of AP-COP (which is used to calculate this measure) has poorer reliability than the RMS of speed of AP-COP [48,49]. As such, if clinicians are interested in using the symmetry index to evaluate the contributions of individual-limbs to balance control in sub-acute stroke, using the speed-based symmetry index that has a lower measurement error and better relative reliability is suggested over the displacement-based symmetry index.

RMS of displacements of COP along the AP and ML axes, and inter-limb cross-correlation demonstrated poor-to-moderate relative reliabilities in the present study. Although current findings for RMS of displacements of COP are in accordance with the previous works in healthy young [38] and older adults [47], they do not agree with the findings of another study in chronic stroke where a greater reliability was shown for RMS of displacements of COP (ICC=0.91) [13]. This finding emphasizes the importance of determining reliability of measures for every distinct population of interest. Low reliability of RMS of COP, inter-limb cross-correlation, and ellipse area suggest that they might not be appropriate choices for characterizing balance in the sub-acute stage of stroke recovery.

In general, the frequency-domain measures included in our study demonstrated poor-to-good reliability. This generally lower reliability of frequency-domain measures than the other force plate-based measures can be due in part to the short sampling duration of the COP time-series [38]. In clinics, it is feasible to capture the COP time-series in trials as short as 30 seconds. However, the frequency content of the signals sampled over shorter durations may differ from those with longer durations [38]. Thus, the longer a signal, the more likely it is to capture low frequency events. A longer data acquisition period might improve the reliability of frequency-domain force plate-based measures. Previous work in healthy older adults showed that to obtain an ICC≥0.9 for the median power frequency of AP-COP and ML-COP, respectively, 20 and 13 repetitions of 30 seconds-long trials must be averaged [47]. Thus, this solution seems less feasible in clinical settings, since fatigue due to the lengthy tests can negatively impact the assessment results.

Our findings showed that sample entropy of displacements of both AP-COP and ML-COP had moderate-to-good relative reliabilities, with a slightly higher measurement error for sample entropy of AP-COP than ML-COP. A similar but slightly higher ICC estimate was observed in chronic stroke for sample entropy calculated from the resultant COP time-series (ICC_3,1_=0.80) [13]. Similar to the frequency-domain measures, the length of the force plate signal is one of the factors that affect the values of sample entropy of COP [50]. Using trials longer than one minute has been recommended for computing sample entropy values [50].

It has been recommended that, for the purpose of decision-making in medical and sport sciences, measures with reliability values above 0.9 should be considered as having high reliability, between 0.9 and 0.8 as having moderate reliability, and less than 0.8 as having insufficient reliability [25,26]. Therefore, our study suggests that dir-WBA, mean speeds of AP and ML-COP, and speed-based symmetry index can be considered as reliable force plate-based measures for clinical evaluation of balance in individuals with sub-acute stroke, since all have both relative reliability values greater than 0.8, and low measurement errors.

This study was not without limitations; we used 30 seconds-long time-series to estimate the reliability of force plate-based measures in quiet standing with eyes open, therefore our findings might not apply to situations where individuals are unable to stand for at least 30 seconds, to other variations of standing, or balance assessments administered in sitting or with eyes-closed. Our reliability values are within-session test-retest reliability estimates; however, it is likely that clinicians evaluate balance control over consecutive days, rather than multiple times within a session, thus establishing between-session reliability of force plate-based measures can be more clinically useful. We identified the reliability values in the sub-acute stage of stroke recovery; therefore, the findings might not be generalizable to the chronic stroke, or to other neurological conditions.

## 5. CONCLUSIONS

Our findings suggest that the directional weight-bearing asymmetry, mean speeds of AP and ML-COP, and speed-based symmetry index, calculated from only one 30 seconds upright standing trial, are reliable force plate-based measures for balance assessments of individuals within the sub-acute stage of stroke recovery. These reliable measures can be used to objectively detect specific balance problems experienced by people in this population. Better balance assessments using objective and reliable measures will lead to better understanding of the extent of balance impairments and will, in turn, results in better rehabilitation interventions. Future research is required to determine validity of these force plate-based balance measures in stroke population.

## Data Availability

The participants of this study did not give written consent for their data to be shared publicly; therefore, supporting/raw data is not available.

## ACKNOWLEDGMENTS

Authors would like to thank Cynthia Danells, Anthony Aqui, James Borrelli, and Christiane Junod for helping with participant recruiting and data acquisition.

## REFERENCES

[1] Foster EJ, Barlas RS, Bettencourt-Silva JH, Clark AB, Metcalf AK, Bowles KM, et al. Long-Term Factors Associated With Falls and Fractures Poststroke. Front Neurol. 2018 Apr 3;9:210. doi:10.3389/fneur.2018.00210

[2] Hyndman D, Ashburn A. People with stroke living in the community: Attention deficits, balance, ADL ability and falls. Disability and Rehabilitation. 2003 Jan;25(15):817–22. doi:10.1080/0963828031000122221

[3] Schmid AA, Wells CK, Concato J, Dallas MI, Lo AC, Nadeau SE, et al. Prevalence, predictors, and outcomes of poststroke falls in acute hospital setting. JRRD. 2010;47(6):553. doi:10.1682/JRRD.2009.08.0133

[4] Teasell R, McRae M, Foley N, Bhardwaj A. The incidence and consequences of falls in stroke patients during inpatient rehabilitation: Factors associated with high risk. Archives of Physical Medicine and Rehabilitation. 2002 Mar;83(3):329–33. doi:10.1053/apmr.2002.29623

[5] Berg K, Wood-Dauphinee S, Williams JI. The Balance Scale: reliability assessment with elderly residents and patients with an acute stroke. Scand J Rehabil Med. 1995 Mar;27(1):27–36. PMID: 7792547

[6] Mansfield A, Inness EL. Force Plate Assessment of Quiet Standing Balance Control: Perspectives on Clinical Application within Stroke Rehabilitation. Rehabilitation Process and Outcome. 2015 Jan;4:RPO.S20363. doi:10.4137/RPO.S20363

[7] McGinnis PQ, Hack LM, Nixon-Cave K, Michlovitz SL. Factors that influence the clinical decision making of physical therapists in choosing a balance assessment approach. Physical Therapy. 2009 Mar;89(3):233-.

[8] Sawacha Z, Carraro E, Contessa P, Guiotto A, Masiero S, Cobelli C. Relationship between clinical and instrumental balance assessments in chronic post-stroke hemiparesis subjects. J NeuroEngineering Rehabil. 2013;10(1):95. doi:10.1186/1743-0003-10-95

[9] Duarte M, Freitas SMSF. Revisão sobre posturografia baseada em plataforma de força para avaliação do equilíbrio. Rev bras fisioter. 2010 Jun;14(3):183–92. doi:10.1590/S1413-35552010000300003

[10] Laroche D, Kubicki A, Stapley PJ, Gremeaux V, Mazalovic K, Maillefert JF, et al. Test– retest reliability and responsiveness of centre of pressure measurements in patients with hip osteoarthritis. Osteoarthritis and Cartilage. 2015 Aug;23(8):1357–66. doi:10.1016/j.joca.2015.03.029

[11] Tamburella F, Scivoletto G, Iosa M, Molinari M. Reliability, validity, and effectiveness of center of pressure parameters in assessing stabilometric platform in subjects with incomplete spinal cord injury: a serial cross-sectional study. J Neuroeng Rehabil. 2014 May 13;11:86. doi:10.1186/1743-0003-11-86

[12] Eng JJ, Chu KS. Reliability and comparison of weight-bearing ability during standing tasks for individuals with chronic stroke. Arch Phys Med Rehabil. 2002 Aug;83(8):1138–44. doi:10.1053/apmr.2002.33644

[13] Jagroop D, Aryan R, Schinkel-Ivy A, Mansfield A. Reliability of unconventional centre of pressure measures of quiet standing balance in people with chronic stroke. Gait & Posture. 2023 May 1;102:159–63. doi:10.1016/j.gaitpost.2023.03.021

[14] Gasq D, Labrunée M, Amarantini D, Dupui P, Montoya R, Marque P. Between-day reliability of centre of pressure measures for balance assessment in hemiplegic stroke patients. J Neuroeng Rehabil. 2014 Mar 21;11:39. doi:10.1186/1743-0003-11-39

[15] Martello SK, Boumer TC, Almeida JC de, Correa KP, Devetak GF, Faucz R, et al. Reliability and minimal detectable change of between-limb synchronization, weight-bearing symmetry, and amplitude of postural sway in individuals with stroke. Research on Biomedical Engineering. 2017 Jun;33(2):113–20. doi:10.1590/2446-4740.06816

[16] Bernhardt J, Hayward KS, Kwakkel G, Ward NS, Wolf SL, Borschmann K, et al. Agreed Definitions and a Shared Vision for New Standards in Stroke Recovery Research: The Stroke Recovery and Rehabilitation Roundtable Taskforce. Neurorehabil Neural Repair. 2017 Sep;31(9):793–9. doi:10.1177/1545968317732668

[17] Gray VL, Ivanova TD, Garland SJ. Reliability of center of pressure measures within and between sessions in individuals post-stroke and healthy controls. Gait & Posture. 2014 May;40(1):198–203. doi:10.1016/j.gaitpost.2014.03.191

[18] Schinkel-Ivy A, Singer JC, Inness EL, Mansfield A. Do quiet standing centre of pressure measures within specific frequencies differ based on ability to recover balance in individuals with stroke? Clinical neurophysiology. 2016;127(6):2463–71. doi:10.1016/j.clinph.2016.02.021

[19] Singer JC, Mochizuki G. Post-Stroke Lower Limb Spasticity Alters the Interlimb Temporal Synchronization of Centre of Pressure Displacements Across Multiple Timescales. IEEE transactions on neural systems and rehabilitation engineering. 2015;23(5):786–95. doi:10.1109/TNSRE.2014.2353636

[20] Maki BE, Holliday PJ, Topper AK. A Prospective Study of Postural Balance and Risk of Falling in An Ambulatory and Independent Elderly Population. J Gerontol. 1994 Mar 1;49(2):M72–84. doi:10.1093/geronj/49.2.M72

[21] Mansfield A, Danells CJ, Zettel JL, Black SE, McIlroy WE. Determinants and consequences for standing balance of spontaneous weight-bearing on the paretic side among individuals with chronic stroke. Gait Posture. 2013 Jul;38(3):428–32. doi:10.1016/j.gaitpost.2013.01.005

[22] Rahimzadeh Khiabani R, Mochizuki G, Ismail F, Boulias C, Phadke CP, Gage WH. Impact of Spasticity on Balance Control during Quiet Standing in Persons after Stroke. Stroke Research and Treatment. 2017;2017:1–10. doi:10.1155/2017/6153714

[23] Kanekar N, Lee YJ, Aruin AS. Frequency analysis approach to study balance control in individuals with multiple sclerosis. Journal of Neuroscience Methods. 2014 Jan;222:91–6. doi:10.1016/j.jneumeth.2013.10.020

[24] Roerdink M, Hlavackova P, Vuillerme N. Center-of-pressure regularity as a marker for attentional investment in postural control: A comparison between sitting and standing postures. Human Movement Science. 2011 Apr;30(2):203–12. doi:10.1016/j.humov.2010.04.005

[25] Vaz S, Falkmer T, Passmore AE, Parsons R, Andreou P. The Case for Using the Repeatability Coefficient When Calculating Test–Retest Reliability. Hempel S, editor. PLoS ONE. 2013 Sep 9;8(9):e73990. doi:10.1371/journal.pone.0073990

[26] Hopkins WG. Measures of reliability in sports medicine and science. Sports Med. 2000 Jul;30(1):1–15.

[27] Borrelli JR, Junod CA, Inness EL, Jones S, Mansfield A, Maki BE. Clinical assessment of reactive balance control in acquired brain injury: A comparison of manual and cable release-from-lean assessment methods. Physiotherapy Research International. 2019;24(4):e1787. doi:https://doi.org/10.1002/pri.1787

[28] Mansfield A, Schinkel-Ivy A, Danells CJ, Aqui A, Aryan R, Biasin L, et al. Does Perturbation Training Prevent Falls after Discharge from Stroke Rehabilitation? A Prospective Cohort Study with Historical Control. Journal of Stroke and Cerebrovascular Diseases. 2017 Oct 1;26(10):2174–80. doi:10.1016/j.jstrokecerebrovasdis.2017.04.041

[29] McIlroy WE, Maki BE. Preferred placement of the feet during quiet stance: development of a standardized foot placement for balance testing. Clin Biomech (Bristol, Avon). 1997 Jan;12(1):66–70. doi:10.1016/s0268-0033(96)00040-x

[30] Stergiou N. Nonlinear Analysis for Human Movement Variability. CRC Press; 2016.

[31] Scoppa F, Capra R, Gallamini M, Shiffer R. Clinical stabilometry standardization. Gait & Posture. 2013 Feb;37(2):290–2. doi:10.1016/j.gaitpost.2012.07.009

[32] Schubert P, Kirchner M, Schmidtbleicher D, Haas CT. About the structure of posturography: Sampling duration, parametrization, focus of attention (part I). JBiSE. 2012;05(09):496–507. doi:10.4236/jbise.2012.59062

[33] Paillard T, Noé F. Techniques and Methods for Testing the Postural Function in Healthy and Pathological Subjects. BioMed Research International. 2015;2015:1–15. doi:10.1155/2015/891390

[34] Berg KO, Maki BE, Williams JI, Holliday PJ, Wood-Dauphinee SL. Clinical and laboratory measures of postural balance in an elderly population. Archives of Physical Medicine and Rehabilitation. 1992 Nov 1;73(11):1073–80. doi:10.5555/uri:pii:000399939290174U

[35] Oliveira LF, Simpson DM, Nadal J. Calculation of area of stabilometric signals using principal component analysis. Physiol Meas. 1996 Nov 1;17(4):305–12. doi:10.1088/0967-3334/17/4/008

[36] Rougier PR, Genthon N. Dynamical assessment of weight-bearing asymmetry during upright quiet stance in humans. Gait & Posture. 2009 Apr;29(3):437–43. doi:10.1016/j.gaitpost.2008.11.001

[37] Winter D, Prince F, Stergiou P, Powell C. Medial-lateral and anterior-posterior motor responses associated with centre of pressure changes in quiet standing. Neuroscience Research Communications. 1993 Jan;12(3):141–8.

[38] Carpenter MG, Frank JS, Winter DA, Peysar GW. Sampling duration effects on centre of pressure summary measures. Gait Posture. 2001 Feb;13(1):35–40. doi:10.1016/s0966-6362(00)00093-x

[39] Genthon N, Rougier P, Gissot AS, Froger J, Pélissier J, Pérennou D. Contribution of Each Lower Limb to Upright Standing in Stroke Patients. Stroke. 2008 Jun;39(6):1793–9. doi:10.1161/STROKEAHA.107.497701

[40] Golomer E, Dupui P, Séréni P, Monod H. The contribution of vision in dynamic spontaneous sways of male classical dancers according to student or professional level. Journal of Physiology-Paris. 1999;93(3):233–7. doi:10.1016/S0928-4257(99)80156-9

[41] Goldberger AL, Amaral LAN, Glass L, Hausdorff JM, Ivanov PCh, Mark RG, et al. PhysioBank, PhysioToolkit, and PhysioNet. Circulation. 2000 Jun 13;101(23):e215–20. doi:10.1161/01.CIR.101.23.e215

[42] Ramdani S, Seigle B, Lagarde J, Bouchara F, Bernard PL. On the use of sample entropy to analyze human postural sway data. Med Eng Phys. 2009 Oct;31(8):1023–31. doi:10.1016/j.medengphy.2009.06.004

[43] Koo TK, Li MY. A Guideline of Selecting and Reporting Intraclass Correlation Coefficients for Reliability Research. J Chiropr Med. 2016 Jun;15(2):155–63. doi:10.1016/j.jcm.2016.02.012

[44] Portney LG, Watkins MP. Foundations of Clinical Research : Applications to Practice. 2nd ed. Upper Saddle River, N.J. : Prentice Hall Health; 2000.

[45] Matheson GJ. We need to talk about reliability: making better use of test-retest studies for study design and interpretation. PeerJ. 2019 May 24;7:e6918. doi:10.7717/peerj.6918

[46] Lexell JE, Downham DY. How to Assess the Reliability of Measurements in Rehabilitation. American Journal of Physical Medicine & Rehabilitation. 2005 Sep;84(9):719–23. doi:10.1097/01.phm.0000176452.17771.20

[47] Lafond D, Corriveau H, Hébert R, Prince F. Intrasession reliability of center of pressure measures of postural steadiness in healthy elderly people. Arch Phys Med Rehabil. 2004 Jun;85(6):896–901. doi:10.1016/j.apmr.2003.08.089

[48] Ruhe A, Fejer R, Walker B. The test–retest reliability of centre of pressure measures in bipedal static task conditions – A systematic review of the literature. Gait & Posture. 2010 Oct;32(4):436–45. doi:10.1016/j.gaitpost.2010.09.012

[49] Jeter PE, Wang J, Gu J, Barry MP, Roach C, Corson M, et al. Intra-session test-retest reliability of magnitude and structure of center of pressure from the Nintendo Wii Balance Board™ for a visually impaired and normally sighted population. Gait & Posture. 2015 Feb;41(2):482–7. doi:10.1016/j.gaitpost.2014.11.012

[50] Montesinos L, Castaldo R, Pecchia L. On the use of approximate entropy and sample entropy with centre of pressure time-series. J NeuroEngineering Rehabil. 2018 Dec;15(1):116. doi:10.1186/s12984-018-0465-9

